# Activating β-catenin pathway mutations predict favorable outcomes after transarterial chemoembolization in unresectable hepatocellular carcinoma

**DOI:** 10.1101/2023.09.28.23296283

**Authors:** Kelley Weinfurtner, Martin Kurian, Daniel Ackerman, Abashai Woodard, Wuyan Li, Jennifer Crainic, Michael C. Soulen, Mandeep Dagli, Susan Shamimi-Noori, Jeffrey Mondschein, S. William Stavropoulos, Shilpa Reddy, Tamim Khaddash, Emma E. Furth, Evan S. Siegelman, Stephen J. Hunt, Gregory J. Nadolski, David E. Kaplan, Terence P.F. Gade

**Affiliations:** Division of Gastroenterology and Hepatology, University of Pennsylvania; Perelman School of Medicine, University of Pennsylvania; Department of Radiology, University of Pennsylvania; Division of Interventional Radiology, University of Pennsylvania; Corporal Michael J Crescenz VA Medical Center, Philadelphia, PA; Department of Pathology and Laboratory Medicine, University of Pennsylvania; Department of Cancer Biology, University of Pennsylvania

**Author notes:** **Corresponding author:** Terence P. F. Gade, Assistant Professor of Radiology and Cancer Biology, University of Pennsylvania Perelman School of Medicine, 652 BRB II/III, 421 Curie Blvd, Philadelphia, PA 19104-6160.

## Abstract

**Background:** Transarterial chemoembolization (TACE) is the most common treatment in unresectable hepatocellular carcinoma (HCC); however, response rates and durability vary widely, and patients frequently require repeat treatments. With the development of alternative locoregional and systemic therapies for patients with HCC, identifying predictors of response to TACE has become increasingly important for a patient population with minimal hepatic reserve. Preliminary data suggests β-catenin pathway mutations may predict favorable response to liver tumors following bland embolization. We hypothesized that activating β-catenin pathway mutations would lead to improved outcomes following TACE in patients with unresectable HCC.

**Material and Methods:** Patients with a clinical diagnosis of HCC planned for TACE were enrolled in a prospective cohort study at two academic medical centers from April 2016 to October 2021. Liver biopsies were taken at time of TACE and mutational profiles determined using a custom next generation sequence panel. Patients with at least one follow-up MRI were included. Primary outcome was objective response rate (ORR) of the targeted tumor at first imaging follow-up. Objective response was defined as complete (CR) or partial response (PR) by mRECIST criteria. Secondary outcomes were CR of targeted tumor at first imaging and at best response, time to target tumor progression (TTP), and overall survival (OS).

**Results:** 53 HCC tumors from 50 patients were included in the analysis. Most patients had BCLC stage B disease (28/53, 52.8%) with underlying cirrhosis (42/53, 84.0%) that was well compensated (45/53 Child-Pugh A, 84.9%). Median size of targeted tumor was 4.0 cm (IQR 2.5-6.3 cm). First follow-up imaging was done at a median of 37 days after TACE (IQR 32-63 days) with ORR of 46/53 (86.8%), including 15 tumors with CR (28.3%). At best response, ORR was 49/53 (92.5%), including 19 tumors with CR (35.9%). 19/53 lesions (35.9%) had progression during the study period with a median TTP of 13.7 months and median OS of 23.4 months. Despite similar ORRs (20/22, 90.2% vs 26/31, 83.8%, p=0.46), tumors with activating β-catenin pathway mutations had increased rates of CR at first imaging (9/22, 40.9% vs 6/31, 19.4%, p=0.09) and at best response (12/22, 54.5% vs 7/31, 22.6%, p=0.02) when compared to tumors without these mutations, as well as longer TTP (median not yet reached vs 8.3 months, p=0.02).

**Conclusions:** In patients with unresectable HCC, activating mutations in β-catenin pathway have better and more durable response to TACE – a finding that may help guide therapeutic decision making in this heterogeneous population.

## INTRODUCTION

Hepatocellular carcinoma (HCC) is the 2^nd^ leading cause of cancer-related death worldwide and fastest growing cause of cancer-related mortality in the United States.^1^ Despite efforts to increase HCC screening, the majority of patients still present with unresectable disease and a median survival less than two years.^2^ This dismal prognosis underscores the limited therapeutic options for this population due to resistance to standard chemotherapies, underlying liver dysfunction, and significant tumor heterogeneity complicating the development of targeted therapeutics.^3^ Further, precision medicine approaches have been hindered by limited access to patient tissue as the diagnosis of HCC can be made from clinical history and imaging alone.

Developed to maximize therapeutic effect in patients with HCC and underlying liver dysfunction, transarterial chemoembolization (TACE) delivers chemotherapy and embolic agents to the tumor by targeting branches of the hepatic artery directly feeding it. By leveraging the liver’s unique dual blood supply, TACE selectively promotes ischemia-induced tumor necrosis while sparing the surrounding liver parenchyma. TACE has been shown to improve survival and is currently the most common treatment in unresectable HCC.^4^ However, this is a very heterogeneous group in regard to tumor burden, tumor biology, and response to TACE. Patients frequently require repeat treatment which can lead to progressive liver dysfunction and death. With the development of other promising therapeutics, including transarterial radioembolization (TARE) and immunotherapy alone or in combination with anti-angiogenic therapy or tyrosine kinase inhibitors, there is a growing need to predict which tumors will respond to TACE to prioritize treatment options in the setting of limited hepatic reserve.

Studies investigating underlying tumor pathogenesis and response to TACE have been lacking, largely due to lack of tissue available for these patients. In a small study that included both primary and metastatic liver tumors, β-catenin pathway mutations were predicative of response to bland transarterial embolization (TAE) with 5/9 (55.6%) responders having CTNNB1 mutations compared to 2/9 (22.2%) non-responders.^6^ Further, the top three mutations predictive of response were CTNNB1 itself and MEN1 and NCOR1, both of which encode proteins involved in Wnt/β-catenin signaling. β-catenin pathway mutations have also been shown to impact response to immunotherapy, namely shorter median progression-free survival (2.0 vs 7.4 months, p<0.0001) and shorter medial overall survival (9.1 vs 15.2 months, 0.11) in patients with unresectable HCC receiving any type of immunotherapy.^7^

In this study, we aimed to determine impact of β-catenin pathway mutations on patient outcomes following TACE from a prospective cohort of patients with unresectable HCC who underwent tumor biopsy for genomic profiling prior to TACE.

## METHODS

### Enrollment

Adult patients with a clinical diagnosis of HCC planned for TACE were recruited from the Interventional Oncology Clinic and multidisciplinary Hepatic Tumor Clinics at two tertiary care academic institutions (Figure 1). Patients were excluded if they (1) were a candidate for resection or orthotopic liver transplantation; (2) did not have an HCC tumor amenable to computed topography (CT) or ultrasound (US)-guided percutaneous biopsy as determined by the treating interventional radiologist; (3) had prior treatment to the target HCC tumor; (4) had an uncorrectable thrombocytopenia/bleeding disorder or anti-coagulation could not be held; (5) had a contraindication to contrast-enhanced magnetic resonance imaging (MRI) for follow-up imaging; or (6) did not have at least one post-treatment MRI. This study was approved by Institutional Review Boards (IRB) at both institutions (University of Pennsylvania IRB #823696 and Corporal Michael J Crescenz VA Medical Center IRB #01779) and all research was performed in accordance with their guidelines/regulations, including obtaining informed consent from all participants.

**Figure 1:**
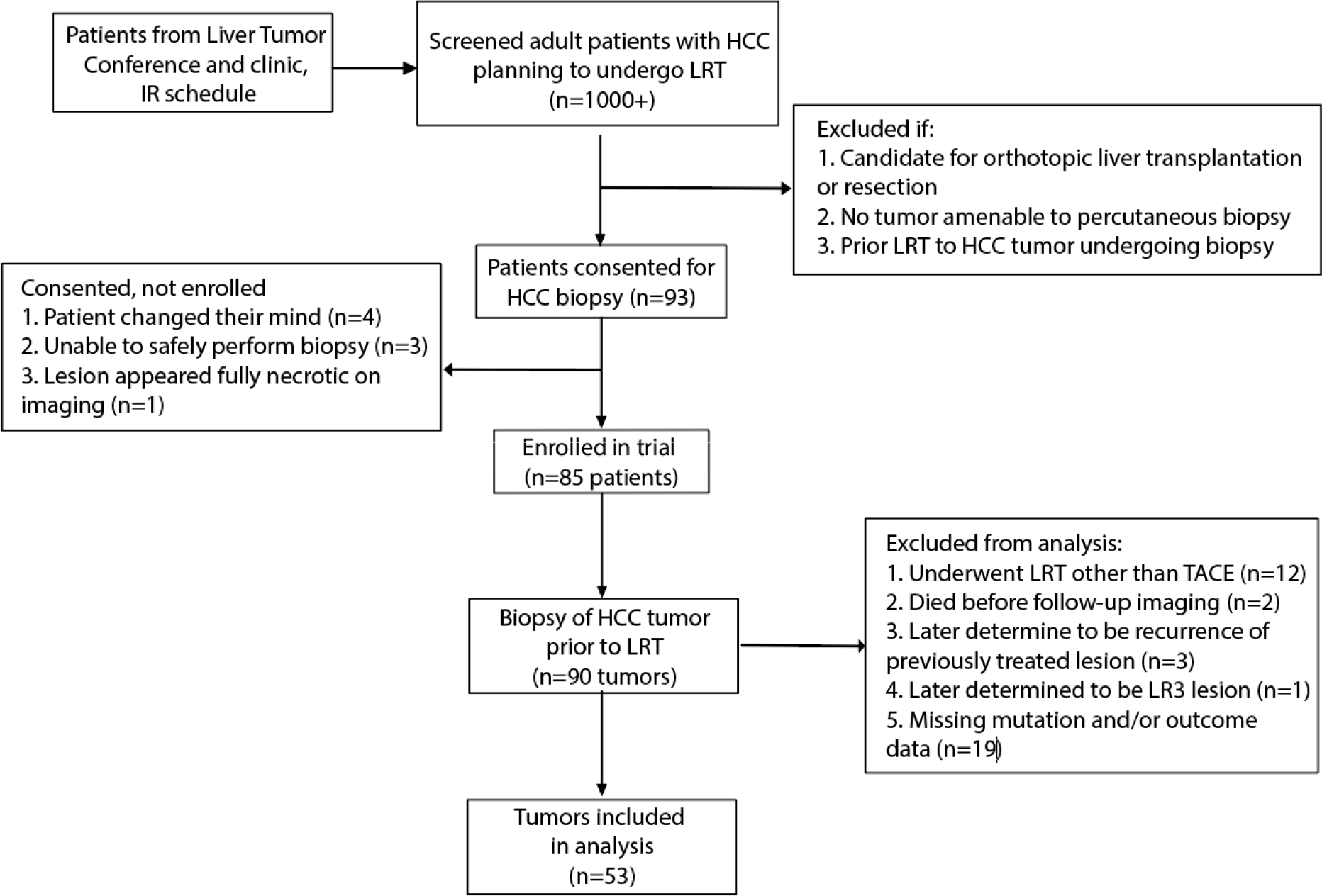
Enrollment Schema

### Biopsy and TACE Procedure

All biopsies were performed by attending interventional radiologists at time of TACE. Briefly, the catheter was positioned into the target vessel and pre-embolization arteriography was performed. Subsequently, the target lesion was identified on US and/or CT. Under imaging guidance, a 17G trocar needle was advanced through the tumor capsule and an 18G BioPince Full Core biopsy needle (Argon Medical, Frisco, TX) was inserted coaxially to obtain core biopsies. TACE procedure***.

### Sample processing

At least two core biopsy samples were sent directly to Surgical Pathology for standard clinical care processing, including formalin fixation and paraffin-embedding with subsequent hematoxylin and eosin staining for histology review. One core biopsy was snap frozen for subsequent DNA isolation. Genomic profiling was done using a custom targeted sequencing amplicon panel developed in collaboration with Swift Biosciences. The assay targeted 234 genomic regions found to be commonly mutated in HCC using publicly available whole exome sequencing data. After generating libraries, samples were multiplexed at 20-30 samples per MiSeq sequencing run. SNVs and indels were called at allele frequencies >5% using the LoFreq variant caller.

### Data collection

All study data was prospectively collected and managed using a Research Electronic Data Capture database. Patient demographics, disease-specific information, and procedure details were obtained from chart review and discussion with the treating interventional radiologist. Histology slides were reviewed by hepatobiliary pathologists (EEF or DJ) with more than 25 years of experience for the presence of malignancy and grade of differentiation. All follow-up cross-sectional imaging was reviewed by a single fellowship-trained body radiologist (ES) with more than 25 years of experience to determine the modified response evaluation criteria in solid tumors (mRECIST) score for the targeted HCC tumor.

### Outcomes and Statistical Analysis

The primary outcome was objective response rate (ORR) of the targeted tumor at first imaging follow-up defined as complete (CR) or partial response (PR) by mRECIST divided by total. The secondary outcomes were (1) CR by mRECIST of targeted tumor at first imaging follow-up; (2) CR by mRECIST of targeted tumor at best response; (3) time to target tumor progression (TTP) defined as time from best response to tumor progression by mRECIST; (4) overall survival (OS) defined as time from enrollment to death or last follow-up. Clinical characteristics and outcomes were compared using Pearson’s chi square, Fisher’s exact, Student’s t-test, Wilcoxon rank-sum, Kruskal-Wallis tests, or Pearson pairwise correlation. Survival analysis was done using Kaplan-Meir log-rank. All data were analyzed using Stata/IC 16.1.

## RESULTS

### Patient and tumor characteristics

50 patients with 53 HCC tumors met inclusion criteria and were included in this analysis. The majority of patients were male (42/50, 84.0%) and white (32/50, 64.0%) with a median age of 64.4 years old (SD 8.1), consistent with the demographics of HCC patients in the United States (Table 1). Most patients had underlying cirrhosis (42/50, 84.0%) that was well compensated (45/53, 84.9% Child-Pugh A, 8/53, 15.1% Child-Pugh B) with median MELD of 8 (IQR 7-10). Median size of targeted tumor was 4.0 cm (IQR 2.5-6.3) with majority in the right lobe only (39/53, 73.6%). The majority of patients had BCLC stage B disease (28/53, 52.8%) with a median of 2 tumors (IQR 1-4) at time of enrollment. On our pathologist’s review of H&E staining, the pathologic diagnosis was well or moderately differentiated HCC in 35/51 tumors (68.6%, 2 tumors with no pathology), poorly differentiated HCC in 4/51 tumors (7.8%), and cholangiocarcinoma (CCA) or combined HCC and CCA (cHCC-CCA) in 5/51 tumors (9.8%). In the other 7 tumors (13.7%), no malignancy was found on pathology. Genomic profiling found activating Wnt pathway mutations in 22/53 tumors (41.5%).

**Table 1.**
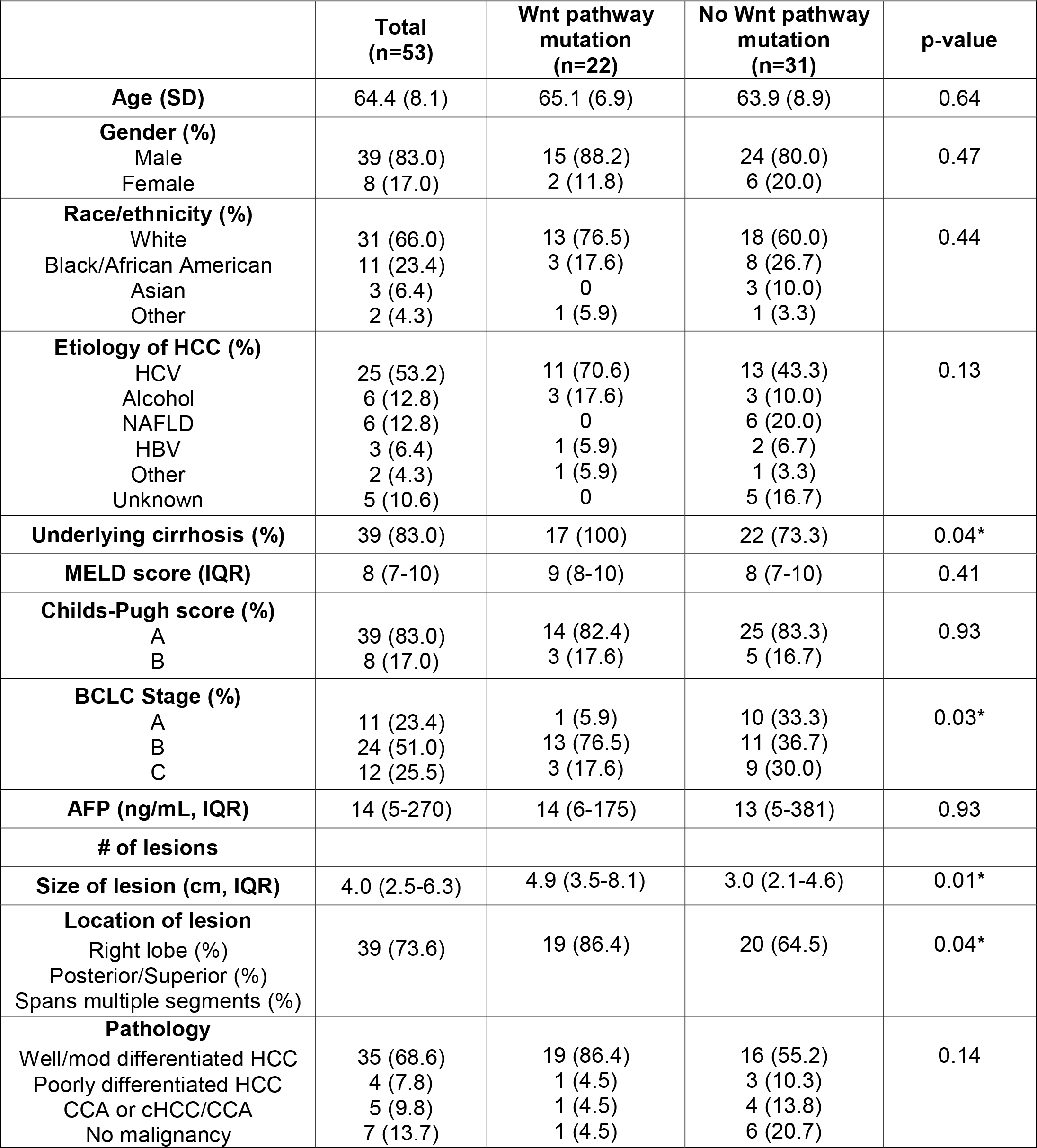
Patient Characteristics.

Patients with β-catenin pathway mutations were more likely to have BCLC stage B disease (17/22, 77.3% vs 11/31, 35.4%, p = 0.006), and larger target tumor size (4.9 cm, IQR 3.5-8.1 cm vs 3.0 cm, IQR 2.1-4.6 cm, p=0.01) than those without these mutations (Table 1). There was also a trend to more patients with HCV (14/22, 73.6% vs 14/31, 45.2%) and fewer with NAFLD (0/22, 0% vs 6/31, 19.4%) or unknown etiology of liver disease (0/22, 0% vs 5/31, 16.1%, overall p=0.11), as well as a trend to more tumors with well/moderately differentiated HCC (19/22, 86.4% vs 16/31, 55.2%) and fewer tumors with poorly differentiated, CCA or cHCC-CCA, or no malignancy on pathology (p=0.14)

### TACE outcomes

The first follow-up imaging after TACE was done at a median of 37 days (IQR 32-63 days) and showed an ORR of 46/53 (86.8%), including 15 tumors with CR (28.3%). At best response, ORR was 49/53 (92.5%), including 19 tumors with CR (35.9%). During the study period, 19/53 tumors (35.9%) had progression with a median TTP of 13.7 months and a median OS of 23.4 months. We found no difference in ORR at first imaging (20/22, 90.9% vs 26/31, 83.9%, p=0.69) nor ORR at best response (21/22, 95.5% vs 28/31, 90.3%, p=0.63) between tumors with β-catenin pathway mutations and those without. However, tumors with β-catenin pathway mutations had increased rates of CR at 1^st^ imaging (9/22, 40.9% vs 6/31, 19.4%, p=0.09) and at best response (12/22, 54.5% vs 7/31, 22.6%, p=0.02) (Figure 2A,B). These tumors also had more durable response to TACE with longer TTP (NYR vs 8.3 months, p=0.02) (Figure 2C).

**Figure 2:**
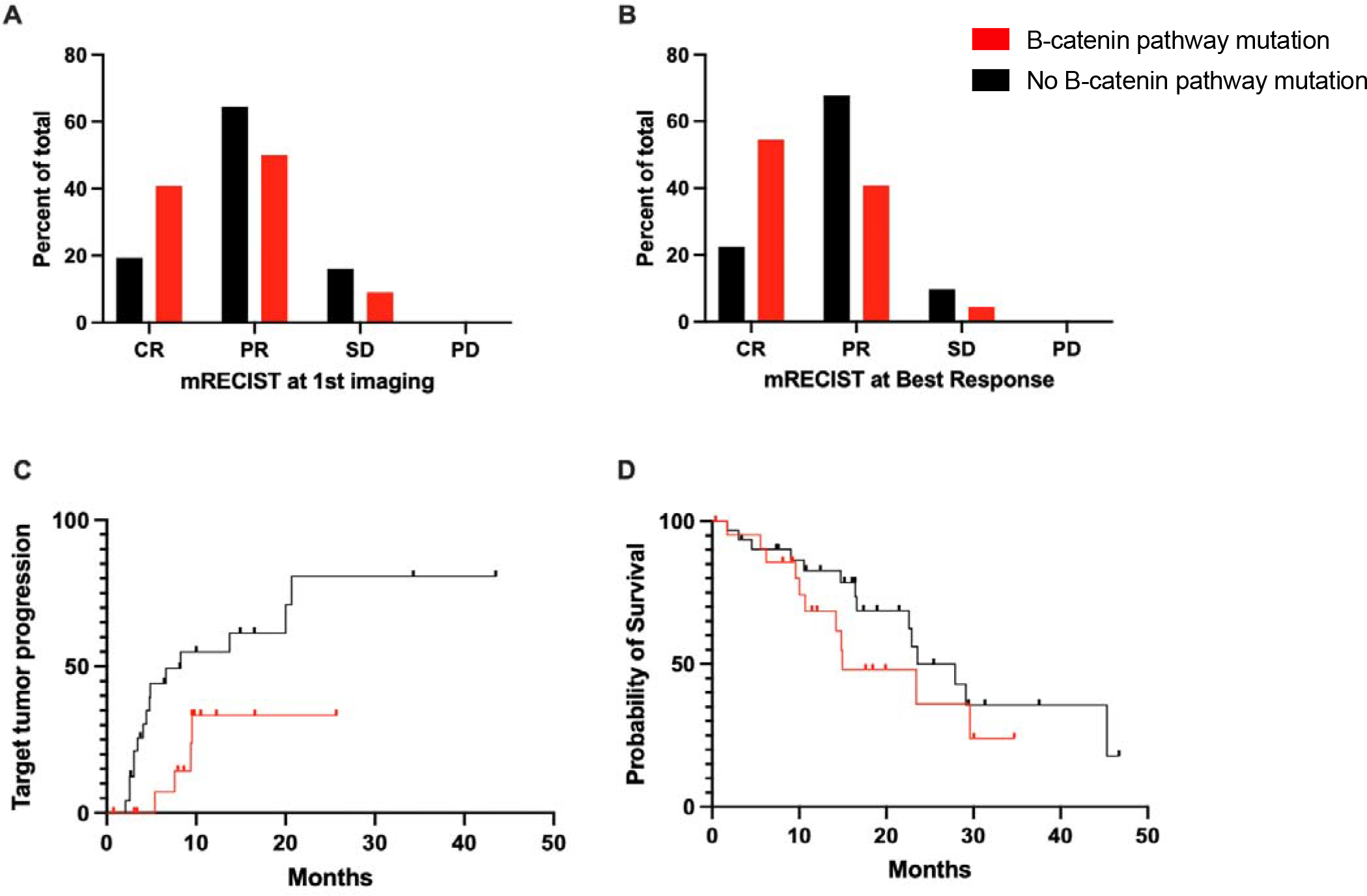
TACE outcomes by mutation status Red represents patients with mutations in B-catenin pathway and black represents patients without mutations in B-catenin pathway. Response following TACE is reported as (A) mRECIST score at first imaging; (B) mRECIST score at best response; (C) time to tumor progression; and (D) overall survival.

There was no difference in OS (15.0 vs 23.6 months, p=0.27) (Figure 2D). There was a trend to shorter TTP in tumors with cHCC-CCA or CCA on pathology (5.7 months vs 13.7 months vs 20.7 months vs 9.4 months in well/moderately differentiates and poorly differentiated, respectively, p=0.29). However, even if we only included tumors with pathology-confirmed HCC, tumors with β-catenin pathway mutations still had higher rates of CR (12/17, 70.6% vs 7/21, 33.3%, p=0.02) and longer TTP (NYR vs 6.6 months, p=0.02).

## DISCUSSION

Over the past two decades, cancer treatments have made significant strides in precision medicine specifically developing or repurposing treatments that target tumors based on the individual patient’s tumor biology to maximize therapeutic benefit while minimizing toxicity. These types of treatment are greatly needed in liver cancer given the high prevalence of underlying liver dysfunction at baseline; however, the field lags behind of other types of cancer as tissue is not required for the diagnosis of HCC. Despite being the most common treatment for patients with unresectable HCC, little is known regarding the impact of tumor biology on response to TACE. In this study, we report patient outcomes following TACE from a prospective cohort of patients with unresectable HCC who underwent a research biopsy prior to TACE and showed that tumors with β-catenin pathway mutations had a more favorable response to TACE with higher rates of CR and longer TTP than tumors without these mutations – a finding that may be a first step toward precision-guided intervention in heterogeneous population.

To the best of our knowledge, this is the largest cohort of HCC tumors prospectively biopsied prior to locoregional therapy, allowing a unique opportunity to study the impact of tumor pathogenesis on TACE outcomes. While several groups have reported on patient and imaging characteristics that predict TACE outcomes, only Ziv et al previously investigated the role of tumor genetics and response to TACE.^cit^ Using a combination of prospectively and retrospectively collected data for 65 patients, Ziv et al demonstrated that tumors with mutations in NRF2 pathway had much shorter time to local progression. They did not report the mutation status for other common driver mutations, including B-catenin pathway proteins; however, in an earlier study that included both primary and metastatic liver tumors, the top three genetic mutations most predictive of sensitivity to TACE were all associated with upregulation of the B-catenin pathway, consistent with our findings. We did not find a difference in ORR as most tumors had at least partial; however, complete response has previously been shown to be a better predictor of long-term response to TACE and that is consistent with our finding of longer time to progression in tumors with B-catenin pathway mutations. Small sample size did not allow for a multivariable model; however, the tumors with B-catenin pathway had improved outcomes despite being larger tumors. The lack of survival benefit seen in this group likely due to multiple factors including less patients with BCLC A disease (6% vs 33%), increased frequency of underlying cirrhosis (100% vs 73%), and potential for differential response to systemic therapies.

Notably, tumor biopsy immediately prior to TACE had an acceptable risk profile with less than 2% major complication rate (3/53, 1.8%, 2 bleeding and 1 seeding events). The one patient who had biopsy track seeing with tumor had CCA on pathology, and the seeding did not impact clinical outcome. Further, these biopsies resulted in other clinically relevant information, specifically 10% of lesions diagnosed as HCC had CCA or cHCC-CCA on pathology review, a finding that has significant prognostic and therapeutic implications. On review of the imaging by our expert body radiologist, only one of these tumors was categorized as LIRADS M (others were LIRADS 5 or TIV). While there were fewer poorly differentiated HCC and CCA or cHCC-CCA tumors in the B-catenin pathway mutation group, tumors with B-catenin pathway mutations still had increased rates of CR and longer TTP compared to those without these mutations even if only biopsy-proven HCC tumors were included, demonstrating our findings were not impacted by the inclusion of these misdiagnosed cHCC-CCA or CCA tumors.

In conclusion, this study was the first to look at the impact of activating β-catenin pathway mutations on TACE outcomes in patients with HCC and demonstrated that the presence of these mutations predicts increased the effectiveness and durability of TACE. These findings need to be validated in larger and prospective interventional studies but suggest that patients could be selected for different treatments based on driver mutations, specifically favoring TACE for patients with β-catenin pathway mutations. Further investigation into the mechanisms behind this susceptibility will allow us to identify mechanism of resistance and develop targeted novel or combination therapies.

## Data Availability

All data produced in the present study are available upon reasonable request to the authors.

## *Abbreviations

AFP: alpha-fetoprotein
BCLC stage: Barcelona clinic liver cancer stage
CT: computed topography
CCA: cholangiocarcinoma
HCC: hepatocellular carcinoma
cHCC-CCA: combined hepatocellular and cholangiocarcinoma
FDG PET: fluorodeoxyglucose positron emission tomography
HCV: hepatitis C virus
HBV: hepatitis B virus
IRB: institutional review board
IQR: interquartile range
LIRADS score: liver reporting and data system score
LRT: locoregional therapy
MELD: model for end-stage liver disease
MRI: magnetic resonance imaging
NAFLD: non-alcoholic fatty liver disease
PDX: patient-derived xenografts
SD: standard deviation
TERT: telomerase reverse transcriptase
US: ultrasound

## Acknowledgements

Research reported in this publication was supported by Penn Center for Precision Medicine Accelerator Grant and VA CSR&D I01 CX-001933.

## *Author contributions

